# Trait mindfulness is associated with less amyloid, tau, and cognitive decline in individuals at risk for Alzheimer’s disease

**DOI:** 10.1101/2021.05.17.21257320

**Authors:** Cherie Strikwerda-Brown, Hazal Ozlen, Alexa Pichet Binette, Marianne Chapleau, Natalie L. Marchant, John C. S. Breitner, Sylvia Villeneuve, for the PREVENT-AD Research Group

## Abstract

**BACKGROUND AND OBJECTIVES:** Mindfulness, defined as non-judgmental awareness of the present moment, has been associated with an array of mental and physical health benefits, including improved cognitive functioning and changes to brain structure and function. Mindfulness may also represent a protective factor for Alzheimer’s disease (AD). Here, we tested the potential protective effect of trait mindfulness on cognitive decline and AD pathology in older adults at risk for AD dementia.

**METHODS:** Two hundred sixty-one nondemented older adults with a family history of AD dementia were included from the PREVENT-AD observational cohort study, including 124 with Aβ- and tau-positron emission tomography (PET) scans. All participants completed measures of trait mindfulness and longitudinal cognitive assessments. Multivariate partial least squares analyses were used to examine relationships between combinations of different facets of trait mindfulness and (1) cognitive decline, (2) Aβ, and (3) tau.

**RESULTS:** Higher levels of mindful nonjudgment, describing, and nonreactivity were associated with less cognitive decline in attention, global cognition, and immediate and delayed memory. Higher levels of mindful nonjudgment and nonreactivity were related to less Aβ-PET signal in bilateral medial and lateral temporoparietal and frontal regions. Higher levels of mindful acting with awareness, describing, nonjudgment, and nonreactivity were associated with less tau-PET signal in bilateral temporal regions, namely entorhinal cortex, amygdala, and parahippocampal, fusiform, and inferior temporal gyri. These relationships remained significant when additional demographic (age, sex, education and APOE status) and psychological (personality traits and neuropsychiatric symptoms) were included in analyses. When mindfulness, personality traits, and neuropsychiatric symptoms were entered into the same model, mindful nonjudgment showed the strongest association with cognitive decline and Aβ.

**DISCUSSION:** Trait mindfulness is associated with less cognitive decline and less Aβ and tau in the brain in older adults at risk for AD dementia. In addition to its other health benefits, trait mindfulness may also represent a psychological protective factor for AD dementia. Longitudinal studies examining the temporal relationship between trait mindfulness and AD markers, along with mindfulness intervention studies with cognition and pathology as outcome variables, will be important for further clarifying the potential protective benefits of mindfulness on AD risk.

## Introduction

Mindfulness is defined as the ability to engage in non-judgmental awareness of the present moment [1]. Growing evidence suggests that both the dispositional trait and the formal practice of mindfulness are associated with an array of mental and physical health benefits, including reduced depression, anxiety, and stress [2], improved sleep [3], less pain [4], and better cardiovascular function [5]. Mindfulness has also been related to changes in brain structure and function [6], along with better cognitive performance [7], including in older adults [8]. The goal of the current study was to examine the possible protective benefits of trait mindfulness on cognitive decline and Alzheimer’s disease (AD) pathology in nondemented older adults at risk of AD dementia.

Mindfulness could be an important factor influencing AD risk given its association with depression, anxiety, sleep disturbance, and cardiovascular health: well-known risk factors for AD dementia [9-12]. Mindfulness is also a strong component of psychological wellbeing, which is increasingly recognised as an important category of behavioural factors protecting against the development of AD dementia [13]. The effects of psychological wellbeing on cognitive decline have been shown to be comparable to that of clinical and lifestyle factors such as diabetes, smoking, and physical inactivity [14].

In the absence of an imminent treatment or cure for AD dementia, increasing focus is being placed on early prevention. Encouragingly, up to 40% of the risk for dementia is attributed to modifiable behavioural factors [15]. It is estimated that even a 10% reduction in these risk factors would reduce the prevalence of AD-related dementia by over 8% [16]. AD is characterised by a long preclinical stage, involving the accumulation of Aβ and tau pathology, along with subtle cognitive decline, up to two decades prior to the onset of dementia [17]. This phase offers a window of opportunity for studying the behavioural factors associated with the earliest stage of AD.

Leveraging a longitudinal cohort sample, we tested the potential protective effect of trait mindfulness on cognitive decline in older adults at risk of AD dementia. We also assessed the association between trait mindfulness and AD pathology, with the hypothesis that mindfulness could reduce dementia risk via by providing resistance against AD pathology. We collected measures of trait mindfulness along with longitudinal cognitive assessments and Aβ-and tau-positron emission tomography (PET) scans in older adults at increased risk of AD. We then used multivariate statistical analyses to examine associations between combinations of different aspects of trait mindfulness and (1) cognitive decline, (2) Aβ, and (3) tau.

## Methods

### Participants and study design

Two hundred and sixty-one participants were included in this study from the PRe-symptomatic EValuation of Experimental or Novel Treatments for AD (PREVENT-AD) cohort. This ongoing longitudinal observational study enrols older adults who are cognitively normal but have a parent or at least 2 siblings diagnosed with AD-like dementia, placing them at increased risk for sporadic AD [18]. Other enrolment criteria include being over 60 years of age (or between 55 and 59 if their age was less than 15 years from their parent’s age of symptom onset), an absence of major neurological and psychiatric diseases, and intact cognition based on the Montreal Cognitive Assessment [MoCA, score above 25; 19] and the Repeatable Battery for the Assessment of Neuropsychological Status [RBANS; 20] at the time of enrolment. Study enrolment spanned from 2012 to 2017, while longitudinal data collection, including annual cognitive assessment with the RBANS, is ongoing. Since 2017, additional measures have been added to the study. In 2017-2019, a subset of participants underwent Aβ and tau PET scans at a single time point, and in 2018 a measure of trait mindfulness was collected (with an additional 16 participants completing it in 2020/2021). All participants included in the current analyses completed the measure of trait mindfulness and at least one annual cognitive assessment (n = 261), with 124 of these individuals also completing Aβ and tau PET scans. Further details of the relative timing of measures are contained within Supplementary Methods. Participants were all cognitively normal at the time of their baseline cognitive assessment and PET scans, though 12 individuals (including 5 PET participants) developed mild cognitive impairment (MCI) prior to completion of the mindfulness questionnaire. Results remain predominantly similar when these participants with MCI are excluded from analyses (see Supplementary Results).

### Mindfulness

The 39-item self-report Five Facet Mindfulness Questionnaire [FFMQ; 21] was collected for all participants (n = 261). The questionnaire was administered via an online platform for most participants, though 10 individuals chose to receive a paper copy in the post, and a further 16 participants completed it over the telephone. The FFMQ contains five subscales measuring different facets of trait mindfulness: Observing (being aware of, and paying attention to, internal and external stimuli and experiences), Describing (verbally labelling and expressing one’s experiences), Acting with awareness (paying conscious attention to what is happening in the present moment, rather than acting automatically), Nonjudgment (not judging one’s own experiences), and Nonreactivity (allowing experiences to arise without reacting to them). Participants were instructed to rate each item based on the extent to which the statements applied to them, on a Likert scale of 1 (Never or Very Rarely True) to 5 (Very Often or Always True). Scores were calculated for each mindfulness subscale, with higher scores reflecting higher levels of mindfulness. The Nonreactivity subscale consists of 7 items, whereas the remaining subscales consist of 8 items. Scores for Nonreactivity therefore ranged from 7 to 35, while the remaining subscale scores ranged from 8 to 40. Missing values were replaced with the mean of the subject’s responses for each subscale. Three participants with >10% of missing values were excluded from analyses. The FFMQ has demonstrated reliability and validity in clinical as well as non-clinical populations [21, 22].

### Cognition

As part of the PREVENT-AD study, participants underwent annual cognitive testing using the RBANS. The RBANS comprises 12 cognitive subtests, which produce five cognitive domain Index scores (Immediate Memory, Attention, Visuospatial Construction, Language, and Delayed Memory), along with a total score reflective of Global Cognition. Scores are standardised to a mean score of 100 with a standard deviation of 15. Longitudinal cognitive assessment (≥ 2 time points) was available for 257 (98.5%) of participants, with a median follow up time of 5 years (range: 1-7 years). The FFMQ was collected a mean of 1429.29 days (i.e., ∼ 3.9 years) after the baseline cognitive assessment (standard deviation = 620.97 days). For further details of the closest cognitive time point to the FFMQ for each participant, see Supplementary Methods.

### Neuroimaging measures

#### Image acquisition

A subset of participants (n = 124) underwent PET imaging using [^18^F]NAV4694 to assess Aβ burden and [^18^F]AV1451 (flortaucipir) for tau deposition. Aβ scans were performed 40 to 70 minutes post-injection (≈6 mCi), and tau scans 80 to 100 minutes post-injection (≈10 mCi). PET images were acquired at the McConnell Brain Imaging Centre at the Montreal Neurological Institute across two consecutive days. T1-weighted structural MRI scans were also collected at the Brain Imaging Centre of the Douglas Mental Health University Institute, with the following parameters: repetition time = 2300ms, echo time = 2.98ms, 176 slices, slice thickness = 1mm. Scans were performed at a single time point, a mean of 435 days prior to the FFMQ (standard deviation = 333.55 days).

#### Image processing

Aβ- and tau-PET scans were preprocessed using a standard pipeline (https://github.com/villeneuvelab/vlpp). Briefly, T1-weighted structural MRI scans were segmented with the Desikan-Killany atlas using FreeSurfer v5.3 [23]. PET images for each participant were then registered to the corresponding T1 scan, masked to exclude signal from the scalp and cerebrospinal fluid, and smoothed with a 6mm^3^ Gaussian kernel. Standardized uptake value ratios (SUVRs) for each of the Desikan-Killaney regions were calculated using reference regions of the whole cerebellum grey matter for Aβ-PET scans [24] and the inferior cerebellar grey matter for tau-PET [25]. SUVRs from regions demonstrated to accumulate Aβ and tau early in the disease course were included in subsequent analyses. For Aβ, this included 38 SUVRs from the 19 bilateral regions that form the global Aβ index, namely lateral and medial prefrontal, parietal, lateral temporal and cingulate cortices [26]. For tau, 14 SUVRs were included, comprising 7 bilateral regions accumulating early tau pathology in preclinical AD [27]. This included regions from Braak stages I (entorhinal cortex), III (amygdala, and fusiform, parahippocampal, and lingual gyri) and temporal regions from Braak stage IV (inferior and middle temporal gyri). Braak stage II (i.e., the hippocampus) was excluded due to nonspecific binding in this region from the choroid plexus [28].

### Statistical analysis

#### Primary analyses

In order to explore the relationships between combinations of mindfulness facets and (1) longitudinal cognition, (2) Aβ, and (3) tau, three separate multivariate partial least squares analyses (PLS) were employed [29] using PLS Software v6.15.1 on MATLAB v2018a. PLS takes two sets of variables (from two matrices) and uncovers linear combinations of these variables that maximally correlate with one another [30]. For all three PLS analyses, the first matrix contained each mindfulness facet (i.e., Observing, Describing, Acting with Awareness, Nonjudgment, and Nonreactivity) in a separate column, with rows representing individual participants. Mindfulness data were z-scored due to the differing ranges of each subscale. The second matrix contained separate columns for each of the dependent variables, namely each of the six cognitive change scores for PLS 1 (see below for details of calculation) or PET SUVRs for each region of interest for PLS 2 and 3 (38 regions for Aβ, 14 regions for tau). Each row represented a single participant.

Prior to conducting the longitudinal cognition PLS, individual cognitive decline slope terms were calculated for each participant using linear mixed-effects models, which are robust to missing and uneven data distribution in longitudinal studies. Using the lme4 v1.1.26 package [31] in R v1.1.463, separate linear mixed-effects models were performed for each of the six cognitive scores as the outcome (Global Cognition, Immediate Memory, Attention, Visuospatial Construction, Language, and Delayed Memory), which included random slopes and intercepts for each subject, and a fixed effect of time as the explanatory variable. The time variable referred to the number of months since the baseline cognitive assessment. Coefficients of individual longitudinal cognitive change for Global Cognition, Immediate Memory, Attention, Visuospatial Construction, Language, and Delayed Memory were extracted from the linear mixed-effects models, which were then included in the second matrix of the longitudinal cognition PLS analysis.

The PLS analyses produced sets of latent variables relating the mindfulness facets with each dependent variable (longitudinal cognition, Aβ, and tau). Each latent variable consisted of a singular value, a vector of weights for each mindfulness facet, and a vector of weights for each dependent variable. Permutation tests were used to identify the significant latent variables (*p* < .05). Then, for each significant latent variable, bootstrap resampling was employed to ascertain the most stable mindfulness subscales and cognitive scores/brain regions contributing to the multivariate relationship. For the mindfulness subscales, the standard error of the resampled distribution was calculated. The mindfulness facets for which the standard error values did not overlap with zero were considered as significantly contributing to the multivariate relationship with the dependent variable. A bootstrap ratio was produced for each of the dependent variables. Cognitive scores/brain regions with bootstrap values > 1.96 were considered as significant contributors to the multivariate relationship with mindfulness. For Aβ and tau PET PLS analyses, bootstrap ratios for each of the Desikan-Killaney regions-of-interest were projected on 2D brain representations using the ggseg package v1.6.01 [32] in R v1.1.463. Finally, the vectors of weights for each mindfulness subscale and dependent variable were multiplied by the original data for these variables for each participant, producing a weighted score of mindfulness facets and a weighted score of longitudinal cognition, Aβ, and tau for each participant. These scores were then used in subsequent linear regression analyses to examine the potential effect of covariates on the multivariate relationship between mindfulness and the dependent variables. In these models, implemented using the lm function in R, the effect of the weighted mindfulness score on predicting the weighted dependent variable score was explored while controlling for age at the time of FFMQ completion, sex, years of education, APOE status (□4 carrier versus noncarrier), and number of days between completion of the FFMQ and the baseline cognitive assessment/PET scans.

#### Complementary analyses

Given we have previously documented relationships in the PREVENT-AD dataset between psychological variables, such as personality and neuropsychiatric factors, and longitudinal cognition, Aβ, and tau [30, 33], we were also interested in examining the potential influence of these other psychological factors on the multivariate relationships between mindfulness and longitudinal cognition, Aβ, and tau. Participants completed questionnaire measures of personality traits (i.e., extraversion, agreeableness, conscientiousness, neuroticism, and openness) and neuropsychiatric symptoms (depression, anxiety, and stress) (see Supplementary Methods for further details). We first performed univariate Pearson correlation analyses to examine associations between mindfulness traits and the other psychological factors. Subsequently, we repeated our main PLS analyses between mindfulness and longitudinal cognition, Aβ, and tau, including these additional psychological variables alongside the mindfulness facets in the first PLS matrix.

### Standard Protocol Approvals, Registrations, and Patient Consents

All participants provided written informed consent, and all research procedures were approved by the Institutional Review Board at McGill University and complied with the ethical principles of the Declaration of Helsinki.

### Data Availability

Data may be made available by request by a qualified investigator.

## Results

Participant characteristics are displayed in Table 1. The PET subsample was representative of the whole cognition sample, except that the PET subsample was slightly older (*p* = 0.02). No differences in trait mindfulness were apparent between the APOE □4 carriers versus noncarriers in either the full sample or PET subsample (all *p* values > .08).

**Table 1.** Demographic, pathological and clinical characteristics of the sample.

|  | **Cognition sample**<br>**(n = 261)** | **PET subsample**<br>**(n = 124)** |
| --- | --- | --- |
| **Demographics** |  |  |
| Age, years | 67.27 (5.24) | 68.60 (4.90) |
| Sex, F:M (% F) | 186:75 (71.3) | 92:32 (74.2) |
| Education, years | 15.50 (3.46) | 15.25 (3.29) |
| APOE ε4 carriers, n (%) | 101 (38.7) | 76 (38.7) |
| MCI at time of FFMQ, n (%) | 12 (4.6) | 5 (4.03) |
| **PET** |  |  |
| Global Aβ SUVR | | 1.34 (0.34) |
| Entorhinal tau SUVR |  | 1.07 (0.14) |
| **Cognition** |  |  |
| MoCA, baseline (out of 30) | 28.11 (1.59) | 28.16 (1.50) |
| RBANS, baseline |  |  |
| Global Cognition | 102.82 (10.32) | 102.73 (10.66) |
| Attention | 107.02 (15.08) | 106.42 (15.41) |
| Visuospatial | 96.39 (13.63) | 96.71 (13.55) |
| Language | 103.75 (9.96) | 104.02 (10.55) |
| Immediate Memory | 102.57 (11.39) | 101.91 (10.67) |
| Delayed Memory | 102.33 (9.09) | 102.48 (9.28) |
| **Mindfulness (FFMQ)** |  |  |
| Observing | 27.55 (6.05) | 28.31 (5.94) |
| Describing | 29.51 (6.04) | 30.28 (5.56) |
| Acting with awareness | 31.01 (5.71) | 31.23 (5.59) |
| Nonjudgment | 30.66 (5.51) | 31.03 (5.53) |
| Nonreactivity | 23.11 (4.75) | 23.63 (4.49) |

Data is presented as mean (standard deviation) unless otherwise specified. Aβ = amyloid $\beta$ ta;
APOE = apolipoprotein E; FFMQ = Five Factor Mindfulness Questionnaire; MCI = Mild
Cognitive Impairment; MoCA = Montreal Cognitive Assessment; PET = Positron Emission
Tomography; RBANS = Repeatable Battery for the Assessment of Neuropsychological Status;
SUVR = standardized uptake value ratio.
*Notes:* Age is calculated at the time of FFMQ completion. APOE $\epsilon$ 4 carriers had at least one copy of the $\epsilon$ 4 allele.

### PLS Analyses

#### 1. Longitudinal cognition

For the PLS analysis between mindfulness and longitudinal cognition, two significant latent variables emerged (*p* < .001, *p* = .004), explaining 68.03% and 20.32% of the PLS variance, respectively. We report the first latent variable here given that it explained the majority of the PLS variance and was replicated in the subset of 124 participants who completed Aβ and tau PET scans (Supplementary Figure 1). Weights and standard errors for the mindfulness facets and bootstrap ratios for the cognitive index slopes for this latent variable are shown in Figure 1. Higher levels of mindful nonjudgment, describing, and nonreactivity were related to less cognitive decline in attention, global cognition, immediate memory and delayed memory (Figure 1A). The correlation between the weighted mindfulness and longitudinal cognitive scores was .24 (*p* < .001), which accounted for 5.4% of the variance in cognitive change. This relationship between mindfulness and longitudinal cognitive decline remained significant when covariates (age, sex, education, APOE status, and time between completion of the FFMQ and baseline RBANS) were included in the model (β = .21, t = 3.55, *p* < .001). The main results from this PLS analysis were also replicated when participants who had MCI at the time of the FFMQ were removed from analyses, with the exception of the immediate and delayed memory scores which did not reach statistical significance (Supplementary Figure 2).

**Figure 1.**
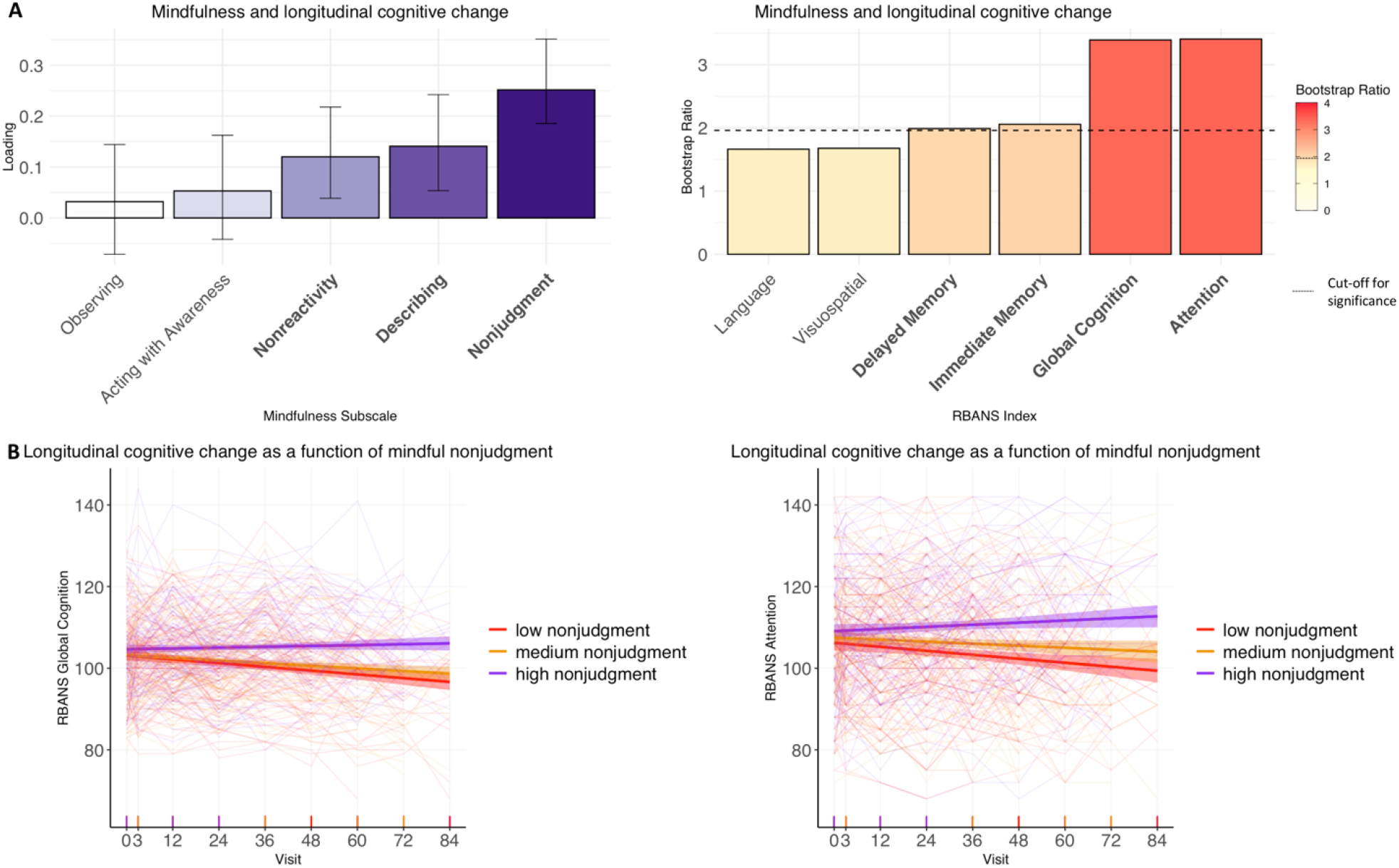
(A) Results of partial least squares (PLS) analyses between mindfulness facets and longitudinal cognitive change. Significant mindfulness facets and cognitive indexes are presented in bold; significant cognition bootstrap ratios are also represented by cognitive indexes with colors above the dotted line on the colorbars. (B) Longitudinal cognitive change for the most robust mindfulness facets and cognitive domains from the PLS analysis, with participants grouped into tertiles of high, medium, and low mindfulness for visualisation purposes.

Figure 1B displays longitudinal cognitive change as a function of mindfulness scores, for the most robust cognitive and mindfulness subscales that contributed to the first latent variable (i.e., longitudinal attention and global cognition as a function of mindful nonjudgment). Participants are grouped into tertiles of high, medium, and low mindfulness for visualisation purposes.

#### 2. Aβ

One significant latent variable emerged for the PLS analysis between mindfulness and Aβ, (*p* = .005), which explained 97.72% of the PLS variance. Loadings and standard errors for the mindfulness facets and bootstrap ratios for the Aβ regional SUVRs that comprise this latent variable are displayed in Figure 2A. Higher levels of mindful nonjudgment and nonreactivity were associated with lower Aβ burden in bilateral medial and lateral temporoparietal and frontal regions (loadings for each brain region of interest are contained within Supplementary Table 1). The correlation between the weighted mindfulness scores and the weighted Aβ regional values was .25 (*p* = .005), accounting for 5.4% of the variance in Aβ. The relationship between mindfulness and Aβ remained significant when covariates (age, sex, education, APOE status, and time between completion of the FFMQ and PET scans) were included in the model (β = .21, t = 2.34, *p* = .02). When participants with MCI at the time of the FFMQ were removed, PLS results remained predominantly similar, though the mindful nonreactivity subscale was no longer significant (Supplementary Figure 3A).

**Figure 2.**
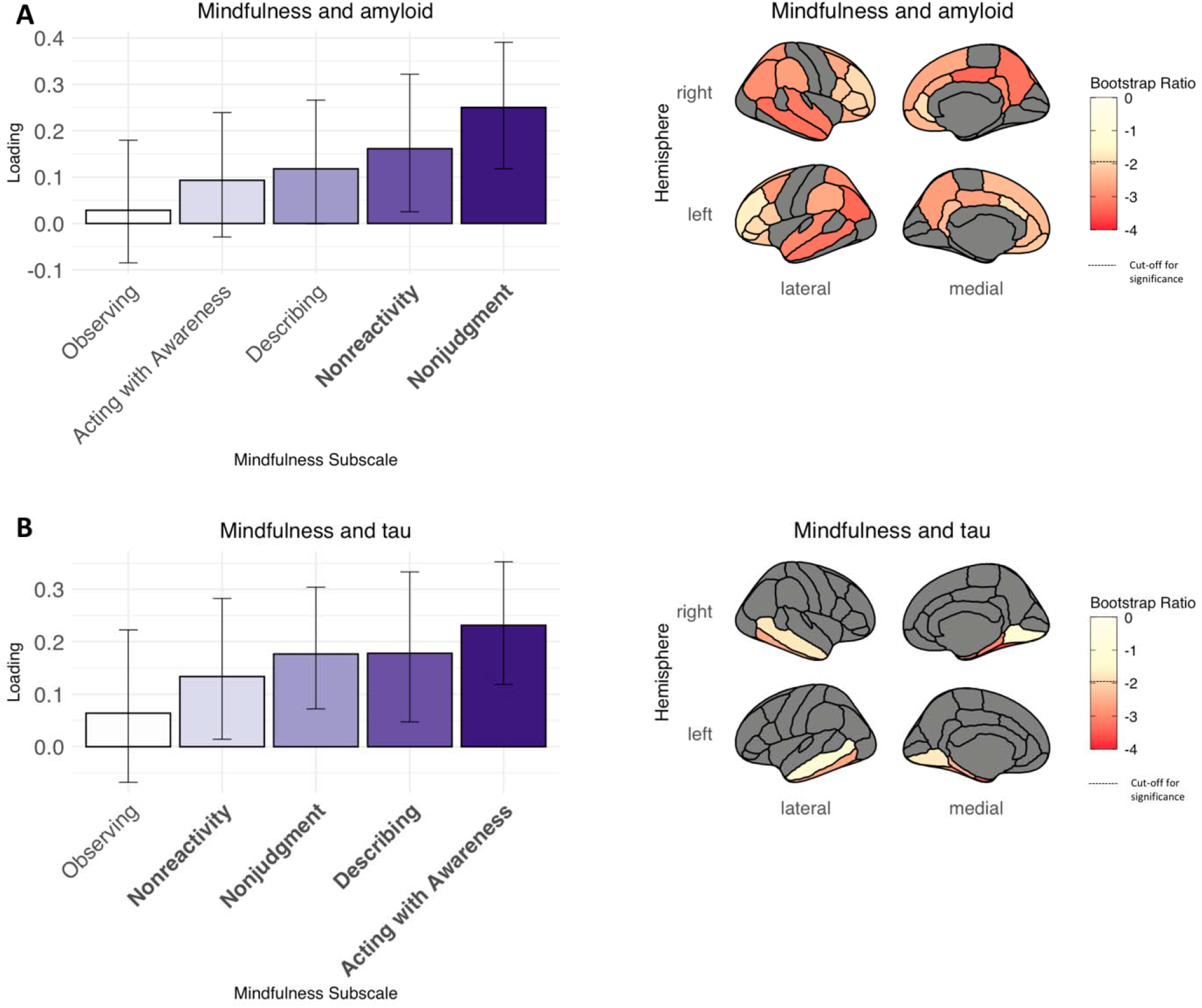
Results of partial least squares (PLS) analyses between mindfulness traits and (A) Aβ and (B) tau SUVRs. Error bars on barplots represent the standard errors. Significant mindfulness facets presented in bold; significant Aβ and tau bootstrap ratios represented by regions with colors below the dotted line on the colorbars. Brain regions shown in grey were not included in analyses.

#### 3. Tau

For the PLS analysis between mindfulness and tau, one significant latent variable emerged (*p* = .007), explaining 92.58% of the PLS variance. Figure 2B shows the loadings and standard error values for the mindfulness facets and bootstrap ratios for the regional tau SUVRs comprising this latent variable. Higher levels of mindful acting with awareness, describing, nonjudgment, and nonreactivity were related to a lower burden of tau in bilateral temporal regions, namely entorhinal cortex, amygdala, and parahippocampal, fusiform, and inferior temporal gyri (see Supplementary Table 1 for the loadings of each region of interest). The correlation between the weighted mindfulness scores and weighted regional tau SUVRs was .26 (*p* = .003), accounting for 6.1% of the tau variance. This relationship between mindfulness and tau remained significant when covariates (age, sex, education, APOE status, and time between completion of the FFMQ and PET scans) were included in the model (β = .25, t = 2.88, *p* = .005). Results of this PLS analysis remained unchanged when participants with MCI at the time of the FFMQ were excluded (Supplementary Figure 3B).

### Complementary analyses

Univariate correlation analyses between mindfulness and other psychological variables revealed that higher levels of mindfulness traits were correlated with less neuroticism, perseverative thinking, anxiety, stress, and depression, and higher conscientiousness, openness, agreeableness, and extraversion (Supplementary Figure 4). When the main PLS analyses between mindfulness and longitudinal cognition, Aβ, and tau, were repeated including these additional psychological variables alongside the mindfulness facets, the majority of the previously uncovered relationships between mindfulness traits and AD markers remained significant, with the exception of the mindful nonreactivity subscale in both the Aβ and tau PLS analyses, which did not reach statistical significance (Supplementary Results; Supplementary Figure 5). This suggests that these relationships are at least partially independent of other psychological factors. For Aβ, the addition of other psychological variables to the PLS analysis also did not explain any additional variance in Aβ compared with the mindfulness traits alone. Importantly, for both the longitudinal cognition (Figure 3A) and Aβ (Figure 3B) PLS analyses, mindful nonjudgment was the strongest contributing psychological variable to the multivariate relationship.

**Figure 3.**
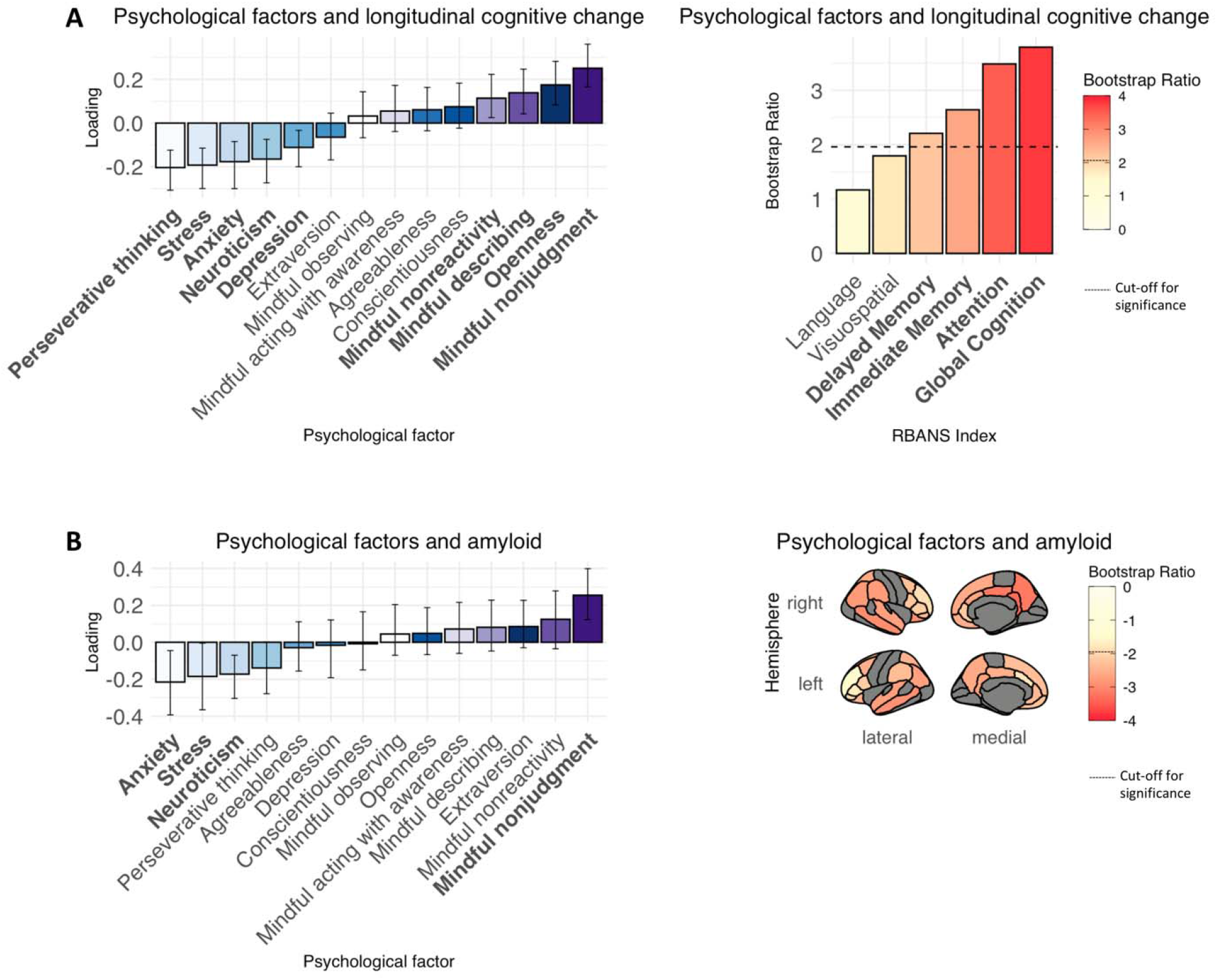
Results of partial least squares (PLS) analyses between psychological variables and (A) longitudinal cognitive change and (B) Aβ SUVR. Error bars on barplots represent the standard errors. Mindfulness variables are presented in shades of purple, remaining psychological factors represented in shades of blue. (A) Significant psychological variables and cognitive indexes presented in bold; significant cognition bootstrap ratios also represented by cognitive indexes with colors above the dotted line on the colorbars. (B) Significant psychological variables presented in bold; significant Aβ bootstrap ratios represented by regions with colors below the dotted line on the colorbars. Brain regions shown in grey were not included in analyses.

## Discussion

The potential health benefits of mindfulness have become increasingly recognised in recent years. In particular, mindfulness is posited to have positive effects on brain health, including improved cognition [7]. Focusing on the preclinical stage of AD, here we explored multivariate relationships between trait mindfulness, cognitive decline, and AD pathology. In older adults at risk for sporadic AD dementia, higher levels of trait mindfulness were related to less cognitive decline, and less Aβ and tau deposition in the brain. Distinct combinations of mindfulness traits were related to cognitive decline, Aβ, and tau, however mindful nonjudgment emerged as a common correlate of all three AD markers. Importantly, while mindfulness was related to other psychological variables including neuropsychiatric symptoms and personality traits, the main associations between mindfulness and cognitive decline, Aβ, and tau remained significant when these other psychological factors were included in analyses. Furthermore, when neuropsychiatric symptoms, personality traits and mindfulness were all entered into the same model, mindful nonjudgment still showed the strongest association with Aβ and cognitive decline. Together, these results highlight a potential role for trait mindfulness as a psychological protective factor for AD.

In older adults at risk of AD dementia, higher levels of trait mindful nonjudgment, describing, and nonreactivity were related to less cognitive decline in global cognition, attention, and immediate and delayed memory over an average of 5 years. The associations between higher levels of mindfulness and less decline in global cognition and attention were also apparent in the subset (249/261) of cognitively unimpaired participants. Mindfulness has previously been associated with better cognitive performance in older adults, particularly in domains of attention and executive function [8]. By definition, attentional control is a primary component of mindfulness, given it involves the directing of attention to one’s present moment experience [1], with improvements in attentional control observed following mindfulness training programs, including in older individuals [34]. Our findings suggest that increased levels of trait mindfulness may also be associated with the maintenance of attentional function over time in cognitively normal older adults at risk for AD dementia, along with other cognitive domains known to be affected in preclinical AD [35]. Further longitudinal studies are required to determine whether older adults who are more mindful in daily life may be able to withstand cognitive decline associated with aging and/or AD, and indeed whether these associations translate to a delaying, or even prevention, of the onset of dementia.

In order to explore whether mindfulness could potentially influence dementia risk via providing resistance against AD pathology, we also examined associations between trait mindfulness and Aβ and tau deposition in the brain. In support of this hypothesis, higher levels of mindful nonjudgment and nonreactivity were related to less Aβ in bilateral medial and lateral temporoparietal and frontal regions, including precuneus, posterior cingulate cortex, and orbitofrontal cortex. These regions are amongst the first cortical areas to accumulate Aβ pathology early in the disease course [26, 36, 37]. It is interesting to note that many of the Aβ regions associated with mindfulness belong to the ‘default mode network’, a widespread functional brain network known to be disrupted in early AD [38]. Mindfulness has also been associated with changes in the activity and functional connectivity of the default mode network, particularly the posterior cingulate [39, 40], which has been proposed as a key candidate mechanism for the effects of mindfulness on psychological wellbeing [41]. Default mode network connectivity therefore represents a potential pathway which mindfulness could be associated with Aβ pathology, which is worthy of further inquiry.

Higher levels of mindful acting with awareness, describing, nonjudgment, and nonreactivity were also related to less tau in regions known to accumulate pathology early in the course of AD, namely entorhinal cortex, amygdala, and parahippocampal, inferior temporal, and fusiform gyri. Medial temporal lobe regions are believed to play important roles in mindfulness, with both the trait and formal practice of mindfulness associated with grey matter volume of the amygdala [42] and hippocampus [43], along with the functional connectivity profiles of these regions [44, 45]. Taken together, the associations uncovered here between mindfulness and Aβ and tau pathology in specific neural regions suggest that future studies examining the potentially mediating role of brain structure and function on these relationships may be of interest.

While distinct combinations of mindfulness traits were associated with cognitive decline, Aβ, and tau, mindful nonjudgment was the only subscale related to all three markers. The associations between mindful nonjudgment, cognition, and AD pathology were highly robust: remaining significant even with the exclusion of MCI participants and the inclusion of other psychological variables in the analyses. Broadly, the construct of mindfulness is considered to incorporate both the attention to present moment experiences, and the attitude of acceptance toward these experiences [46]. The mindful nonjudgment subscale of the FFMQ captures this latter component, namely the tendency to refrain from judging one’s own thoughts and feelings as they arise, via items such as ‘I judge my thoughts as good or bad’ and ‘I criticize myself for having irrational emotions and thoughts’ (both reverse-scored). Mindful nonjudgment has been proposed to play a critical role in reducing stress [47], alleviating symptoms of psychopathology [48] and improving cardiovascular health [49], over and above present moment awareness. Our findings suggest that mindful nonjudgment may also be particularly associated with AD markers.

Preliminary work has begun to explore the effect of mindfulness-based interventions in MCI and early-stage AD dementia. Early evidence from pilot studies suggests that mindfulness interventions are feasible and tolerable in cognitively impaired populations [50], and associated with improvements in mood and quality of life [51, 52]. Their effects on cognition, however, remain less clear, with one study reporting significant cognitive improvements in MCI participants following an eight-week mindfulness intervention [53], though only trend-level differences from pre-to post-intervention were observed in another [52]. While these initial findings are promising, there remains a need for large, well-controlled studies of mindfulness interventions in early AD, with longer follow-up periods [see 54]. These interventions may also be most effective if targeted at earlier, preclinical disease stages [e.g., 55, 56, 57], prior to the onset of extensive and likely irreversible AD pathology, neural damage, and cognitive impairment [17]. Importantly, self-reported mindfulness, as measured by the FFMQ, consistently increases following mindfulness intervention programs [58], including in MCI participants [53], suggesting that this trait is indeed modifiable. Results from the current study point to an association between trait mindfulness and markers of AD in the preclinical stage. Whether mindfulness interventions in this early disease stage may lead to slower rates of AD pathology accumulation and cognitive decline will form a key question for future research.

There are numerous potential psychological, behavioural, and physiological mechanisms by which mindfulness may be associated with AD pathology and cognitive decline. For example, positive impacts of mindfulness on reducing depression, anxiety, stress [2], and modifying personality traits such as neuroticism [59] have been observed. These psychological factors are also associated with increased risk for AD dementia [9, 10], and higher levels of AD pathology and cognitive decline in preclinical disease stages [30, 60, 61]. One common substrate proposed to underlie these psychological factors, and the risk they confer for AD, is the tendency to engage in perseverative negative thinking [33, 62, 63]. Interestingly, mindfulness interventions are believed to also exert their positive effect on mental health via reducing tendencies to engage in perseverative negative thinking [64]. It is therefore possible that the associations between mindfulness and AD may be partially attributable to reductions in perseverative negative thinking and associated psychological symptoms. In the current study, mindfulness traits were associated with perseverative thinking, depression, anxiety, stress, and personality traits in the expected directions. Crucially, however, the main associations between mindfulness and cognition and mindfulness and AD pathology remained apparent even when these other psychological traits were included in analyses. In fact, for both the longitudinal cognition and Aβ analyses, mindful nonjudgment was the strongest contributing psychological variable to the multivariate relationship. Accordingly, the associations between mindfulness and AD markers are unlikely to be fully explained by its indirect effects on other psychological variables.

Other mechanisms are therefore likely involved in the relationships between mindfulness and AD markers. Mindfulness has been associated with an array of physical health benefits, involving similar pathways that are proposed to be involved in AD. Poor cardiovascular health is one of the most significant category of risk factors for the development of AD pathology and its associated dementia syndrome [15, 65, 66]. Mindfulness has also been linked to an array of benefits on vascular health, including blood pressure, heart rate variability, and glucose regulation [5]. These effects may be at least partly attributable to the influence of mindfulness on positive health behaviours such as physical activity, smoking cessation, diet, and sleep [5, 67]. Additional physical health markers such as inflammation, physiological stress (including cortisol), immune system dynamics, and cellular aging, have also been associated with both mindfulness [68-71] and AD [72-75]. More research is required to further explore the relative contribution of each of these potential pathways to the association between mindfulness and early markers of AD.

Overall, higher levels of trait mindfulness were associated with better cognition over time and less Aβ and tau in the brain in older adults at risk for AD dementia. Our multivariate approach permitted in-depth profiling of associations between specific combinations of mindfulness facets and AD markers, though such relationships were also apparent using standard univariate analyses (see Supplementary Table 2). The current findings raise the possibility that cultivating mindfulness in daily life may be a way to reduce vulnerability to AD. As our study was correlational, however, and the participants were approaching the expected age of disease onset, it is also possible that lower mindfulness represents an early symptom of AD. Other limitations of our study include the cross-sectional nature of the PET scans, which were only conducted in a subset of participants. Accordingly, large-scale, longitudinal studies examining the temporal relationship between trait mindfulness and AD markers, along with mindfulness intervention studies with cognition and pathology as outcome variables, will be of utmost importance in refining our understanding of the potential protective benefits of mindfulness for AD.

## Data Availability

Data will be released upon reasonable request

## Acknowledgements

The authors would like to thank the PREVENT-AD participants for volunteering their time for the study. CSB was supported by a Strauss Postdoctoral Fellowship from McGill University. APB was supported by a joint scholarship from the Alzheimer’s Society Canada and the Fonds de Recherche du Québec-Santé. NM was supported by a Senior Fellowship from the Alzheimer’s Society (AS-SF-15b-002). SV was supported by a Canada Research Chair and a Canada Fund for Innovation grant. The PREVENT-AD PET scans were funded by Canadian Institutes of Health Research foundation grant, an Alzheimer’s Association grant, a joint Alzheimer’s Society Canada and Brain Canada Research grant, and a Lemaire foundation donation to SV. The PREVENT-AD cohort was funded by generous support from McGill University, the government of Canada, an unrestricted gift from Pfizer Canada, the Canada Fund for Innovation, the Douglas Hospital Research Centre, the Levesque Foundation, and Genome Quebec Innovation Center. Data used in preparation of this article were obtained from the Pre-symptomatic Evaluation of Novel or Experimental Treatments for Alzheimer’s Disease (PREVENT-AD) program (https:douglas.research.mcgill.ca/stop-ad-centre). A complete listing of the PREVENT-AD Research Group can be found in the PREVENT-AD database: https://preventad.loris.ca/acknowledgements/acknowledgements.php?date=[2018-12-17]. Funding sources had no role in the design and conduct of the study; collection, management, analysis, and interpretation of the data; preparation, review, or approval of the manuscript; and the decision to submit the manuscript for publication.

