## Supplementary for "Trait mindfulness is associated with less amyloid, tau, and cognitive decline in individuals at risk for Alzheimer’s disease"

Perry Pavilion Room E3417.1

6875 Boulevard LaSalle	Montreal, QC

Canada H4H 1R3

**Supplementary Methods**

*Relative timing of measures*

Study enrolment for the PREVENT-AD study spanned from 2012 to 2017, while the FFMQ was added to the study in 2018 (with an additional 16 participants completing it in 2020/2021). The longitudinal time point at which the FFMQ was administered therefore varied between participants (baseline: 1.9%, 12-month: 10.7%, 24-month: 13.0%, 36-month: 13.8%, 48-month: 18.8%, 60-month: 20.3%, 72-month: 18.8, 84-month: 2.7% of participants).

*Additional psychological variables*

In addition to the FFMQ, participants also completed measures of neuropsychiatric symptoms and personality traits. This included the Geriatric Depression Screening Scale (Yesavage et al., 1982), the Geriatric Anxiety Inventory (Pachana et al., 2007), the Stress subscale of the Depression Anxiety and Stress Scales (Lovibond & Lovibond, 1995), the Perseverative Thinking Questionnaire (Ehring et al., 2011), and the Big-5 Personality Inventory (capturing extraversion, agreeableness, conscientiousness, neuroticism, and openness) (John, Donahue, & Kentle, 1991). One hundred and fifty-seven participants (120 with PET scans) had a complete psychological dataset available, including the FFMQ along with these additional psychological questionnaires. The majority of participants (95.3%) completed the FFMQ and additional psychological measures at the same time point (2018), with the remaining participants completing the other psychological measures in 2016 or 2017.

**Results**

*PLS between mindfulness and longitudinal cognition in PET subsample*

As a sensitivity analysis, the PLS analysis between mindfulness and longitudinal cognition was repeated the subset of 124 participants who completed Aβ and tau PET scans. One significant latent variable emerged (*p* < .001), explaining 73.32% of the PLS variance. Weights and standard errors for the mindfulness facets and bootstrap ratios for the cognitive index slopes that comprise this latent variable are shown in Supplementary Figure 1. Higher levels of mindful nonjudgment and describing were related to less cognitive decline in global cognition, immediate memory, delayed memory, and attention. The correlation between the weighted mindfulness and longitudinal cognitive scores was .39 (*p* < .001), which accounted for 14.4% of the variance in cognitive change. This relationship between mindfulness and longitudinal cognitive decline remained significant when covariates (age, sex, education, APOE status, and time between completion of the FFMQ and baseline RBANS) were included in the model (β = .34, t = 3.93, *p* < .001).

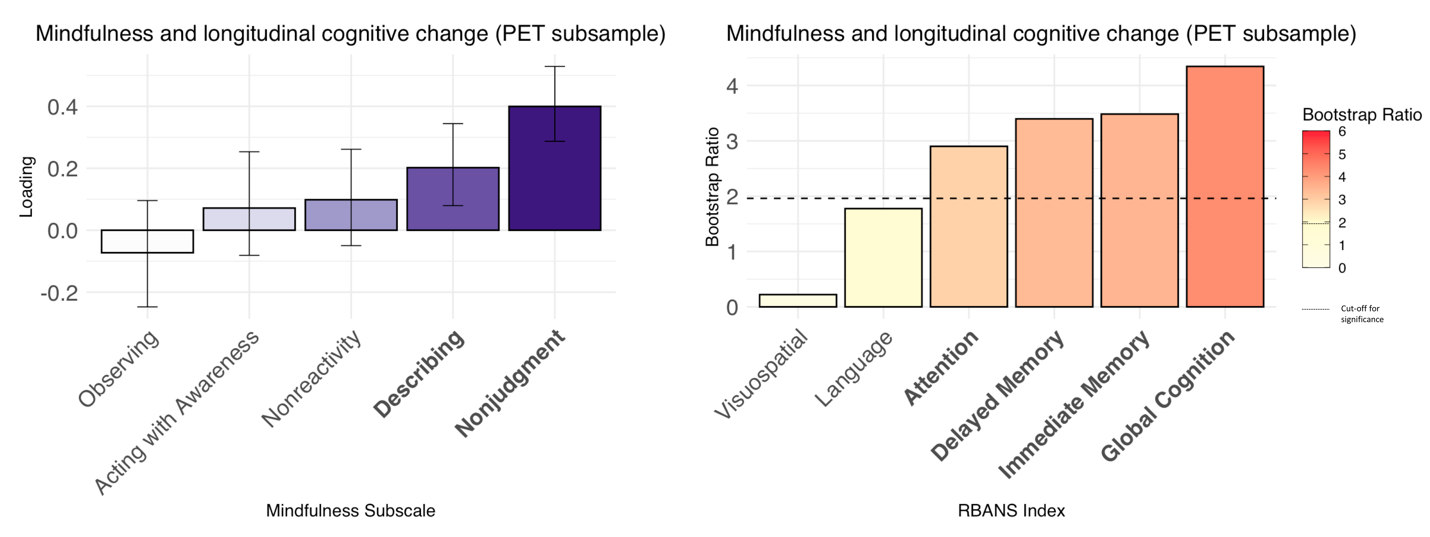

*Supplementary Figure 1.* Results of partial least squares (PLS) analyses between mindfulness facets and longitudinal cognitive change, in the PET subsample (n = 124). Significant mindfulness facets and cognitive indexes are presented in bold; significant cognition bootstrap ratios are also represented by cognitive indexes with colors above the dotted line on the colorbars.

*PLS between mindfulness and longitudinal cognition excluding MCI participants*

The PLS analysis between mindfulness and longitudinal cognition was repeated after removing the 12 participants who had MCI at the time of completion of the FFMQ. Two significant latent variables emerged (*p* < .001, *p* = .008), explaining 60.47% and 25.36% of the PLS variance, respectively. Weights and standard errors for the mindfulness facets and bootstrap ratios for the cognitive index slopes for the first latent variable are shown in Supplementary Figure 2. Higher levels of mindful nonjudgment, describing, and nonreactivity were related to less cognitive decline in attention and global cognition. The correlation between the weighted mindfulness and longitudinal cognitive scores was .24 (*p* < .001), which accounted for 5.2% of the variance in cognitive change. This relationship between mindfulness and longitudinal cognitive decline remained significant when covariates (age, sex, education, APOE status, and time between completion of the FFMQ and baseline RBANS) were included in the model (β = .22, t = 3.50, *p* = .001).

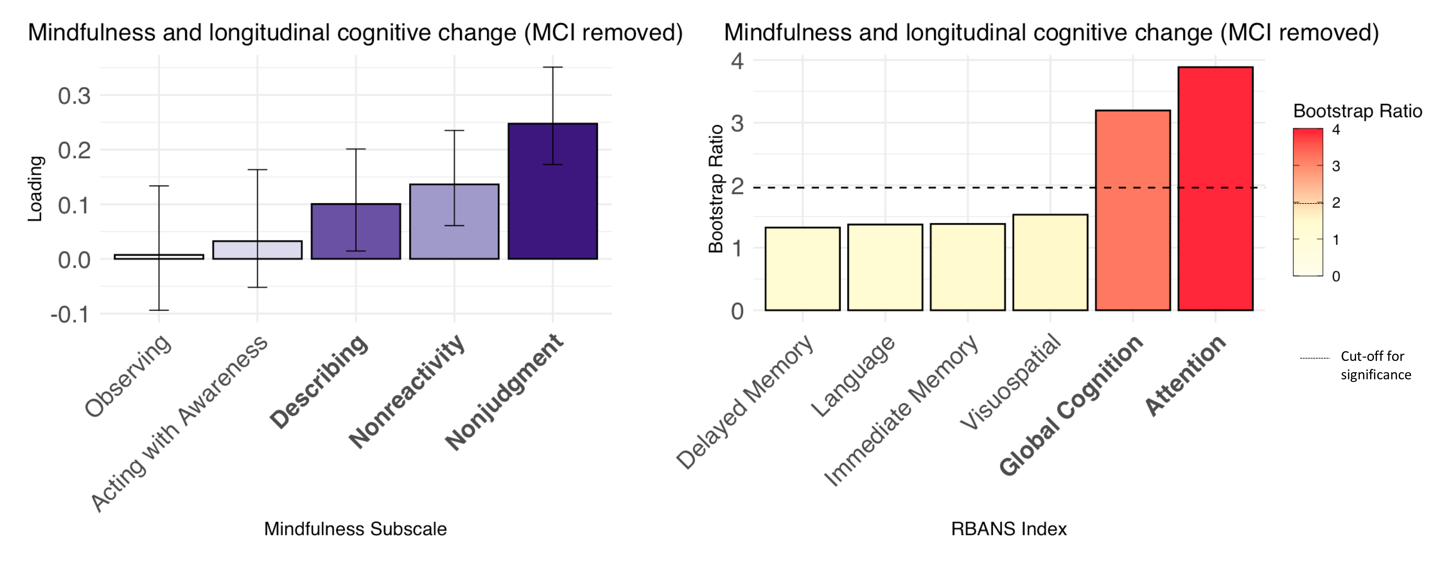

*Supplementary Figure 2.* Results of partial least squares (PLS) analyses between mindfulness facets and longitudinal cognitive change, with the 12 participants who had MCI at the time of completing the FFMQ removed from analyses. Significant mindfulness facets and cognitive indexes are presented in bold; significant cognition bootstrap ratios are also represented by cognitive indexes with colors above the dotted line on the colorbars.

*PLS between mindfulness and Aβ excluding MCI participants*

The PLS analysis between mindfulness and Aβ was repeated after removing the 5 PET participants who had MCI at the time of completion of the FFMQ. One significant latent variable emerged (*p* = .006), which explained 96.98% of the PLS variance. Loadings and standard errors for the mindfulness facets and bootstrap ratios for the Aβ regional SUVRs that comprise this latent variable are displayed in Supplementary Figure 3A. Higher levels of mindful nonjudgment were associated with lower Aβ burden in bilateral medial and lateral temporoparietal and frontal regions (see Supplementary Table 1 for loadings for each brain region of interest). The correlation between the weighted mindfulness scores and the weighted Aβ regional values was .24 (*p* = .008), accounting for 5.0 % of the variance in Aβ. The relationship between mindfulness and Aβ remained significant when covariates (age, sex, education, APOE status, and time between completion of the FFMQ and PET scans) were included in the model (β = .20, t = 2.24, *p* = .03).

*PLS between mindfulness and tau excluding MCI participants*

When the PLS analysis between mindfulness and tau was repeated after removing the 5 MCI PET participants, one significant latent variable emerged (*p* = .02), explaining 91.18% of the PLS variance. Supplementary Figure 3B shows the loadings and standard error values for the mindfulness facets and bootstrap ratios for the regional tau SUVRs comprising this latent variable. Higher levels of mindful acting with awareness, describing, nonjudgment, and nonreactivity were related to a lower burden of tau in bilateral temporal regions, namely entorhinal cortex, amygdala, and parahippocampal, fusiform, and inferior temporal gyri (see Supplementary Table 1 for the loadings of each region of interest). The correlation between the weighted mindfulness scores and weighted regional tau SUVRs was .24 (*p* = .009), accounting for 4.8% of the tau variance. This relationship between mindfulness and tau remained significant when covariates (age, sex, education, APOE status, and time between completion of the FFMQ and PET scans) were included in the model (β = .22, t = 2.54, *p* = .01).

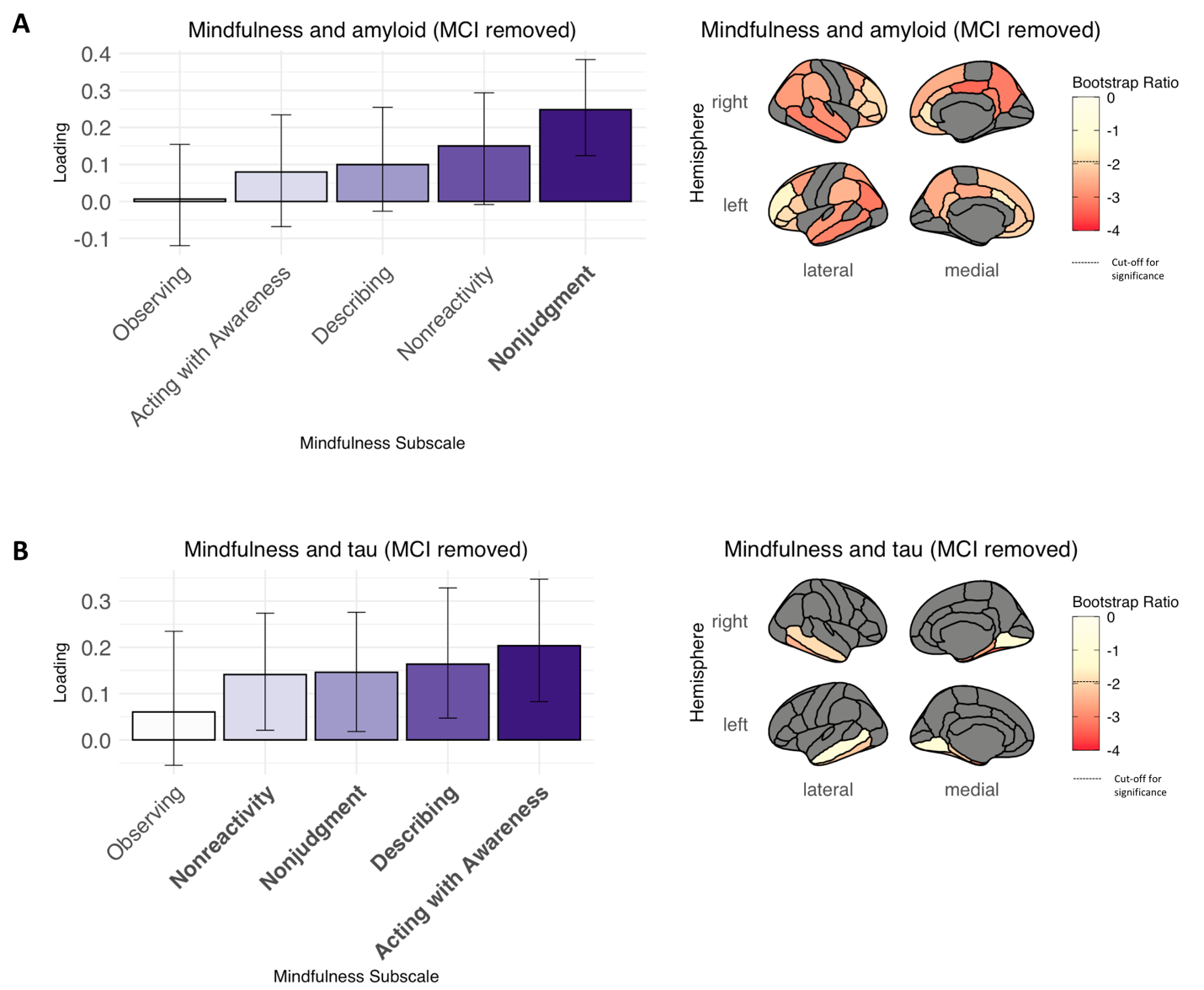
*Supplementary Figure 3.* Results of partial least squares (PLS) analyses between mindfulness traits and (A) Aβ and (B) tau SUVRs, with the 5 PET participants who had MCI at the time of completing the FFMQ removed from analyses. Error bars on barplots represent the standard errors. Significant mindfulness facets presented in bold; significant Aβ and tau bootstrap ratios represented by regions with colors below the dotted line on the colorbars. Brain regions shown in grey were not included in analyses.

*Associations between mindfulness and other psychological variables*

To examine the relationship between mindfulness and other psychological variables, we performed univariate Pearson’s correlations between the 5 mindfulness subscales, and psychological factors of depression, anxiety, stress, perseverative thinking, and the Big-5 personality traits (extraversion, agreeableness, conscientiousness, neuroticism, and openness). Intercorrelations between the measures are displayed in Supplementary Figure 4. Overall, higher levels of mindfulness traits were associated with less neuroticism, perseverative thinking, anxiety, stress, and depression, and higher conscientiousness, openness, agreeableness, and extraversion.

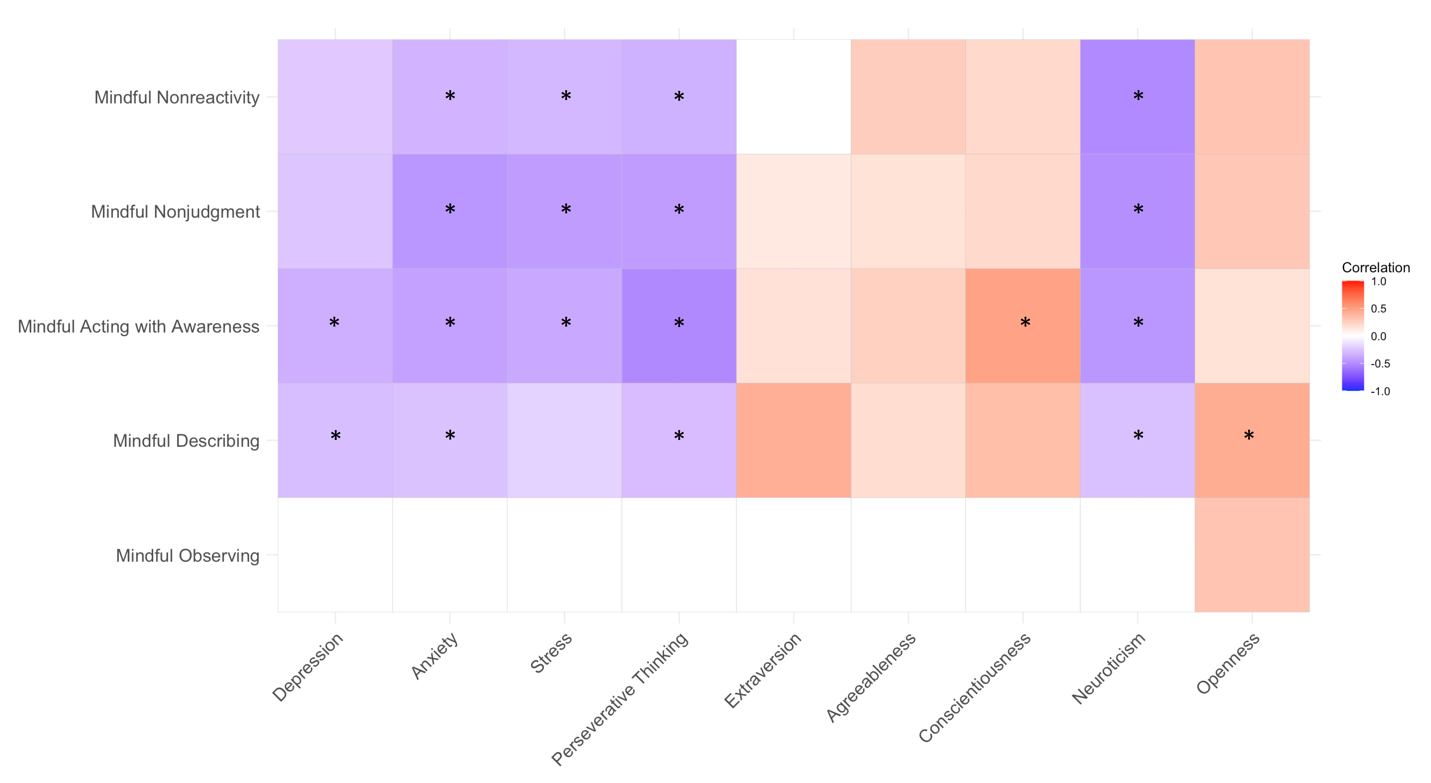

*Supplementary Figure 4.* Correlations between mindfulness facets and psychological factors. Coloured squares represent significant correlations at *p* < .05; * denotes significant correlations at the Bonferoni corrected alpha level of *p* < .0011.

*PLS between psychological variables (including mindfulness) and longitudinal cognition*

For the PLS analysis between psychological factors (including mindfulness) and longitudinal cognition, one significant latent variable emerged (*p* < .001), explaining 76.94% of the PLS variance. Weights and standard errors for the psychological variables and bootstrap ratios for the cognitive index slopes for this latent variable are shown in Figure 3A in the main text. Higher levels of mindful nonjudgment, openness, mindful describing, and mindful nonreactivity, and lower levels of perseverative thinking, stress, anxiety, neuroticism, and depression were related to less cognitive decline in global cognition, attention, immediate memory and delayed memory. The correlation between the weighted psychological and longitudinal cognitive scores was .27 (*p* < .001), which accounted for 6.6% of the variance in cognitive change. This relationship between psychological factors and longitudinal cognitive decline remained significant when covariates were controlled for (β = .24, t = 4.06, *p* < .001).

*PLS between psychological variables (including mindfulness) and Aβ*

One significant latent variable emerged for the PLS analysis between psychological factors and Aβ, (*p* = .005), which explained 95.20% of the PLS variance. Loadings and standard errors for the psychological variables and bootstrap ratios for the Aβ regional SUVRs that comprise this latent variable are displayed in Figure 3B in the main text. Higher levels of mindful nonjudgment lower levels of anxiety, stress, and neuroticism were associated with lower Aβ burden in bilateral medial and lateral temporoparietal and frontal regions (loadings for each brain region of interest are contained within Supplementary Table 1). The correlation between the weighted psychological scores and the weighted Aβ regional values was .25 (*p* = .005), accounting for 5.4% of the variance in Aβ. This relationship between psychological factors and Aβ remained significant when covariates were controlled for (β = .20, t = 2.26, *p* =.03).

*PLS between psychological variables (including mindfulness) and tau*

For the PLS analysis between psychological factors and tau, one significant latent variable emerged (*p* < .001), explaining 91.76% of the PLS variance. Supplementary Figure 5 shows the loadings and standard error values for the psychological variables and bootstrap ratios for the regional tau SUVRs comprising this latent variable. Higher levels of mindful acting with awareness, extraversion, mindful nonjudgment, conscientiousness, mindful describing, and openness, and lower levels of perseverative thinking, stress, anxiety, neuroticism, and depression were related to a lower burden of tau in bilateral temporal regions, namely entorhinal cortex, amygdala, and parahippocampal, fusiform, and inferior temporal gyri, along with the bilateral lingual gyri (see Supplementary Table 1 for the loadings of each region of interest). The correlation between the weighted psychological scores and weighted regional tau SUVRs was .34 (*p* < .001), accounting for 11.0% of the tau variance. This relationship between psychological factors and tau remained significant when covariates were controlled for (β = .31, t = 3.59, *p* <.001).

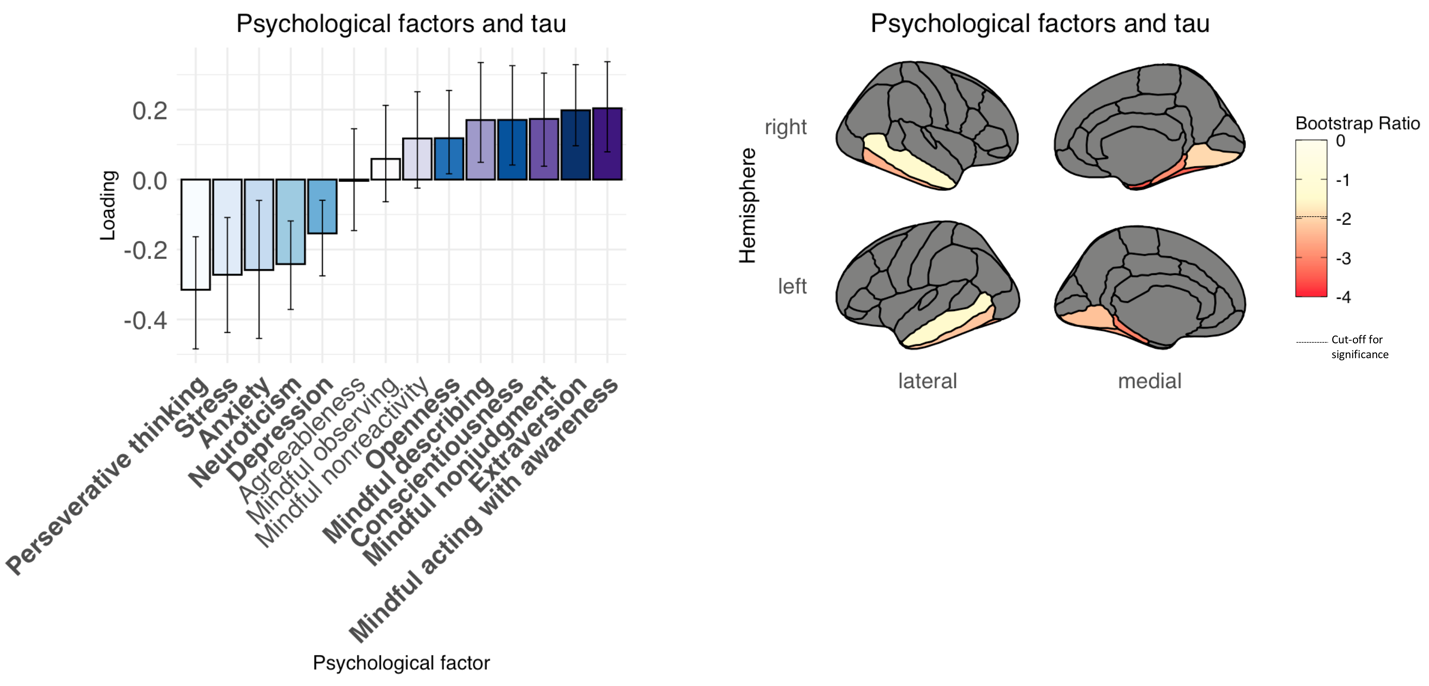

*Supplementary Figure 5.* Results of partial least squares (PLS) analyses between psychological variables and au SUVR. Error bars on barplots represent the standard errors. Mindfulness variables are presented in shades of purple, remaining psychological factors represented in shades of blue. Significant psychological variables presented in bold; significant tau bootstrap ratios represented by regions with colors below the dotted line on the colorbars. Brain regions shown in grey were not included in analyses.

**Supplementary Table 1. Bootstrap ratios for the Aβ and tau SUVRs for each brain region of interest in PLS analyses**

| **Bootstrap ratios** | | | |
| --- | --- | --- | --- |
|  | Mindfulness PLS analysis | Mindfulness PLS analysis (excluding MCI participants) | Psychological factors PLS analysis |
| *Aβ* |  |  |  |
| lh_caudalanteriorcingulate | -1.8734919 | -1.6907783 | -1.7783194 |
| lh_caudalmiddlefrontal | **-2.7959857** | **-2.7653687** | **-2.5168381** |
| lh_inferiorparietal | **-3.3544531** | **-3.1982908** | **-2.702152** |
| lh_isthmuscingulate | **-2.3564787** | **-2.2484739** | **-2.652245** |
| lh_lateralorbitofrontal | **-2.2239437** | **-2.1813409** | **-2.4519329** |
| lh_medialorbitofrontal | **-2.1595068** | **-2.0173254** | **-2.234849** |
| lh_middletemporal | **-3.2191842** | **-3.0830312** | **-2.8850985** |
| lh_parsopercularis | **-2.1501827** | **-2.1297884** | **-2.1135221** |
| lh_parsorbitalis | -1.8352122 | -1.8852723 | -1.9439701 |
| lh_parstriangularis | -1.9419416 | -1.8875855 | -1.698873 |
| lh_posteriorcingulate | **-2.6777596** | **-2.5910454** | **-2.5181627** |
| lh_precuneus | **-2.7518997** | **-2.5593379** | **-2.5915163** |
| lh_rostralanteriorcingulate | **-2.3499436** | **-2.1569107** | **-2.3778415** |
| lh_rostralmiddlefrontal | -1.6353338 | -1.5616708 | -1.5068717 |
| lh_superiorfrontal | **-2.3598802** | **-2.2425971** | **-2.317349** |
| lh_superiorparietal | **-2.8973198** | **-2.7240317** | **-2.4604571** |
| lh_superiortemporal | **-3.0069571** | **-2.8330319** | **-2.7080467** |
| lh_supramarginal | **-2.6144183** | **-2.4570451** | **-2.3487473** |
| lh_frontalpole | -1.7090111 | -1.5674796 | -1.4870107 |
| rh_caudalanteriorcingulate | **-2.5921447** | **-2.4176712** | **-2.6406097** |
| rh_caudalmiddlefrontal | **-2.6016526** | **-2.6018553** | **-2.3000405** |
| rh_inferiorparietal | **-2.9327846** | **-2.7581952** | **-2.8053379** |
| rh_isthmuscingulate | **-3.0114179** | **-3.0431240** | **-3.3295028** |
| rh_lateralorbitofrontal | **-2.5842707** | **-2.5289502** | **-2.9223228** |
| rh_medialorbitofrontal | **-2.4109931** | **-2.3239636** | **-2.5276651** |
| rh_middletemporal | **-3.2754438** | **-3.1039951** | **-2.9548843** |
| rh_parsopercularis | **-2.2400701** | **-2.1443353** | **-2.243993** |
| rh_parsorbitalis | **-2.2280765** | **-2.1740270** | **-2.3588319** |
| rh_parstriangularis | **-2.0311997** | **-2.0176539** | **-2.1265001** |
| rh_posteriorcingulate | **-3.3718445** | **-3.3522258** | **-3.1654589** |
| rh_precuneus | **-3.2386096** | **-3.1457849** | **-3.1747196** |
| rh_rostralanteriorcingulate | -1.8718724 | -1.7532827 | **-2.1629136** |
| rh_rostralmiddlefrontal | -1.9356371 | -1.9331313 | -1.8867705 |
| rh_superiorfrontal | **-2.6118999** | **-2.5432134** | **-2.5896807** |
| rh_superiorparietal | **-2.8695695** | **-2.7754633** | **-2.5903232** |
| rh_superiortemporal | **-3.1687315** | **-2.9163775** | **-2.7213528** |
| rh_supramarginal | **-2.7168398** | **-2.5730250** | **-2.7732291** |
| rh_frontalpole | -1.9020945 | -1.8350009 | -1.7926614 |
| *Tau* |  |  |  |
| lh_amygdala | **-2.6619563** | **-2.3111882** | **-3.9729049** |
| rh_amygdala | **-2.6753075** | **-2.3320525** | **-3.6275580** |
| lh_entorhinal | **-3.2326422** | **-3.0106235** | **-4.0724297** |
| rh_entorhinal | **-3.0205894** | **-2.7808876** | **-3.5841184** |
| lh_fusiform | **-2.7979784** | **-2.1995370** | **-2.6532466** |
| rh_fusiform | **-3.7468009** | **-3.3064363** | **-3.4397013** |
| lh_inferiortemporal | **-2.6174881** | **-2.1668935** | **-2.2124739** |
| rh_inferiortemporal | **-2.5805502** | **-2.4692836** | **-2.5063171** |
| lh_lingual | -1.7199879 | -1.1373202 | **-2.2844222** |
| rh_lingual | -1.2674454 | -1.1187031 | **-1.961354** |
| rh_middletemporal | -1.7642419 | -1.9230721 | -1.4994943 |
| lh_middletemporal | -1.2420421 | -0.95184380 | -1.5173234 |
| lh_parahippocampal | **-2.4271584** | **-2.1211045** | **-3.070771** |
| rh_parahippocampal | **-2.9700458** | **-2.6274183** | **-2.9449368** |

*Note:* Bolded values represent regions with significant bootstrap ratios in the respective PLS analyses, with a cut-off absolute value of 1.96.

**Supplementary Table 2. Pearson’s correlations between mindfulness traits and standard univariate markers of cognitive decline, Aβ, and tau**

| Mindfulness Subscale | Global cognitive decline | Global *Aβ* index | Entorhinal cortex tau |
| --- | --- | --- | --- |
| Observing | -.04 | -.03 | -.14 |
| Describing | .11 | -.12 | **-.20*** |
| Acting with Awareness | .05 | -.10 | **-.18*** |
| Nonjudgment | **-.24***** | **-.26**** | **-.22*** |
| Nonreactivity | .09 | -.17 | -.16 |

*Note:* Bolded values represent significant associations, * *p* < .05, ** *p* < .01, *** *p* < .001
